# “Around the clock”. Exploring health care professionals’ experience of discharge of older people during out of hours from the emergency department: a qualitative study

**DOI:** 10.1101/2024.11.06.24316820

**Authors:** Mary Dunnion, Debbie Goode, Assumpta Ryan, Sonja McIlfatrick

## Abstract

**Background:** Older adults constitute a considerable number of attendances at emergency departments (EDs). Whilst many require hospital admission, a greater focus now is on admission avoidance with older adults being subsequently discharged from EDs. Little is known, however, about the experiences out of hours (OOH) when specialist older person support services are unavailable.

**Aim:** To explore senior health care professionals’ experiences of discharging older people during out of hours from EDs.

**Method:** A qualitative study involving individual semi-structured interviews was used to explore the experiences of healthcare professionals. Fourteen participants in total responded from a wide geographical spread. Data analysis was undertaken using Braun and Clarke’s (2022) six-step framework for Thematic Analysis.

**Results:** One overarching theme was identified focusing on risks and safety for the older person whilst being discharged OOH from the ED. Three sub-themes included “Should they stay, or should they go?”, “Bright lights and noise”, and “New ways of working”, which included risks in discharging an older person OOH from the ED, risks in delaying discharge, and recommendations for future practice. Significant differences were reported between office hours and OOH discharge of older adults from the ED. Diversity in practice assessments, and resources available was also evident across EDs. Significant adverse consequences were identified for older adults having to stay for prolonged periods in the ED.

**Conclusions:** There have been many welcome developments in healthcare services for older people who attend and are subsequently discharged from the ED. However, further innovative practice and collaboration with senior decision makers is needed to meet the healthcare needs of a rapidly ageing population. Safe, equitable and effective discharge practice 24/7 should be a norm for all older people in every ED regardless of location. Gaps in care identified must be addressed taking cognisance of the relevant recommendations for future practice.

## Introduction

Countries worldwide are witnessing an unprecedented population increase in both the number and the proportion of older people (1). The United Nations General Assembly (2) projected that between 2019 and 2030, the number of persons aged 60 years or over will grow by 38 per cent. According to Eliopoulos (3), older adults are generally defined as 65 years of age and older; however, several categories are also recognised, including, young old (65 to 74 years); old (75 to 84 years) and oldest-old (85 years and over) taking cognisance of the diversity between these age groups. Alongside the demographic growth, a current focus on healthy ageing is advocated (4). However, by nature many older adults become unwell which often necessitates a visit to an emergency department (ED). This can often be a difficult experience for some older people who attend the ED with an atypical presentation and in an acutely unwell state. According to Cassarino et al (5) and Corcoran et al (6) attendance at the ED can result in high levels of adverse outcomes for older people. Issues with communication, privacy, and personal care for older people in the ED have been highlighted (7), whilst the ED has been reported as a complex, stressful and unpredictable area (8; 9). Although some older adults may require subsequent admission to hospital from the ED (10), many are discharged home from the department. Whilst the latter option may be the most preferable for older people, Cetin-Sahin (11) and Gettel et al (12) reported on several transitional care needs for patients following discharge.

It is imperative that the discharge process from the ED is safe, comprehensive, effective, and timely. However, outside of office hours (after 5pm until 8am and at all weekends and public holidays) many older person specialist teams are inaccessible in the ED or in the community which poses the question as to the experiences and risks for this cohort being discharged late at night or at weekends (13). Whilst some attention has been given to the discharge of older people from hospital at night (14), a review of the literature has identified a dearth of research regarding out of hours (OOH) discharge of older people from the ED. The aim of this paper was to explore the experiences of Health Care Professionals (HCP) regarding the discharge of older people OOH versus during office hours from the ED.

## Methodology

### Aim and Objectives

This paper was part of an overall study exploring the discharge of older adults out of hours from the ED. This paper focused on the experiences of senior Health Care Professionals’ when discharging older people during OOH versus office hours from the ED.

Objectives:

- To determine the experiences of HCPs when discharging older people from the ED
- To examine the differing practices, if any, between OOH and office hours discharge of older adults from the ED
- To identify recommendations for future practice when discharging older people OOH from the ED

### Study design

In this study a qualitative methodology was used consisting of semi-structured interviews. The questions for the interviews were constructed by the researchers following a review of relevant literature, taking cognisance of the Integrated Care Theoretical Framework (15), the biopsychosocial aspects of health, and the Integrated Care Programmes for Older People (ICPOP). ICPOP was introduced in Ireland with an aim to improve quality of life for older people and to shift from acute hospital, episodic care to community based longitudinal, coordinated, and integrated care (16). This links in with the aim, objectives and theoretical framework of this study. The interview schedule (see Appendix 1) was developed by the authors and reviewed by an ED expert group. A pilot study with one participant was also undertaken to ensure further rigour of the semi-structured questions.

### Population and Recruitment

This study was undertaken in Ireland. Inclusion criteria for participants stipulated that they were required to be senior healthcare professionals working in an ED. Fourteen participants in total responded which included clinical nurse managers, consultants in emergency medicine, advanced nurse practitioners, clinical nurse specialists in frailty, clinical education facilitators and physiotherapists. Participants included a broad range of healthcare professionals and originated from a diversity of urban and rural geographical areas and models of hospitals which added to the rigour of the study.

### Ethical approval

Ethical approval was obtained from Ulster University Research Ethics Filter committee (Project Number: FCNUR-22-017) as well as from the individual hospital Research Ethics Committees (RECs). No national ethics committee currently exists in Ireland; therefore, approval had to be sought from individual hospital RECs. In many of the sites, the Consultant in Emergency Medicine acted as gatekeeper for the research participants as stipulated by the hospital REC. Following confirmation of ethical approval, assistance to identify and invite participants to take part in the study was provided by members of The National Clinical Programme for Emergency Medicine and The Irish Association for Emergency Medicine. All participants were provided with a participation information sheet providing the purpose, risks, benefits of the study, the right to withdraw, and the assurance of anonymity. This was explained by one of the research team to each of the respondents and a consent form was subsequently signed by participants prior to commencement. Consent was also obtained for the possible dissemination of the study findings. Due to the small number of urban and rural EDs in Ireland, it was vital not to breach General Data Protection Regulations (GDPR) by identifying any of the hospitals and/or the participants who contributed to the study. Therefore, careful selection and reporting of the narrative exemplars was undertaken to take cognisance of and avoid the risk of this possibility.

### Data collection

The recruitment period for this study was between February 17^th^ and November 24^th^, 2023. One-to-one interviews were conducted. Written informed consent was obtained by the researcher and consent was reconfirmed at commencement of the interviews with all the participants. All interviews took place via video conferencing and at a time to suit the participants following individual consent to use this option. Due to the busyness of the ED environment, time constraints for participants and the distance between each site for the researcher, this method was deemed to be the most appropriate. Interviews lasted 50 to 60 minutes in duration and were recorded using a digital recorder and via the computer to reduce the risk of loss of the interview data. Data saturation was reached following the final interview as no new data was noted and recruitment then ceased. The recordings were transcribed by one member of the research team and checked for accuracy prior to analysis (MD).

### Data analysis

Data analysis was undertaken using Braun and Clarke’s (17) six-step framework for Thematic Analysis (TA) which included familiarisation with the data set; coding; generating initial themes; developing and reviewing themes; refining, defining, and naming themes; and writing up. TA is a highly effective method to use when looking to analyse and understand a collection of experiences, thoughts, or behaviours (18), therefore taking cognisance of the aim of this study, this framework was selected as appropriate to use in this data analysis. These six steps involved a collaborative approach with the four members of the research team (MD, AR, DG, SMcI). Several meetings took place to interpret the data, identify categories, codes, provisional themes, edit and refine themes to ensure rigour, consensus, and a richer interpretation of the data. These collaborative meetings with the experienced research team at all stages of the study are advocated to ensure reflexive practices in qualitative research (19; 20). The Standards for Reporting Qualitative Research (SRQR) (21) checklist was also completed to ensure rigour and a transparent reporting of the research process (see Appendix 2).

### Findings

Following data analysis, several themes and sub-themes were identified. One recurrent theme highlighted the issues of risks and safety for older people being discharged from the ED OOH. In addition to this over-arching theme, several sub-themes were also identified (see Table 1). Significant differences were reported by the respondents between office hours and OOH discharge of older adults from the ED.

**Table 1:**
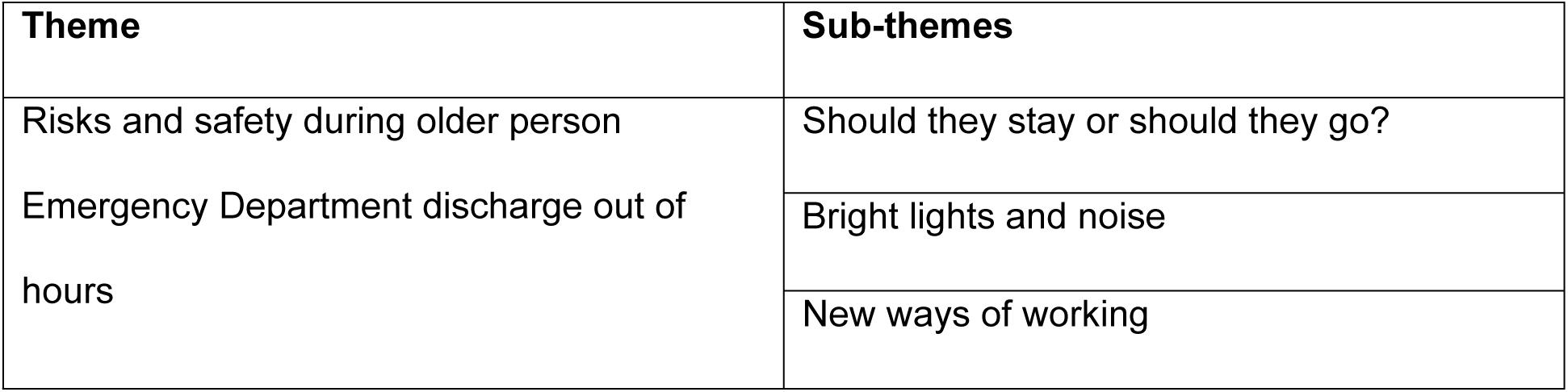
Risks and Safety Theme and Sub-themes.

### Should they stay or should they go?

Participants reported on the risks in OOH ED discharge for the older person. The safety decision to discharge an older adult OOH from the ED or to detain them until reviewed by specialist older people services during office hours was influenced by several factors, including the time of discharge and distance to home. However, if there was any concern that the older person required more support following discharge OOH, it was reported that s/he would be detained or “boarded for review” in ED until 8am when older person specialist staff are available. This was despite staff being aware of the pressures of not having people lying on trolleys.

> “It’s two or four in the morning, but they are a very vulnerable high-risk patient …. and they’re going to go home to an empty house where there is no one. There’s not a light on… we would keep them until the morning.” (Clinical Nurse Manager No. 2)

Participants reported being conscious of the need to ensure safe discharge and refer older people to the specialist supports in the ED and the community OOH. They also reported the benefits of having these specialist older person teams as a great form of support. However, participants also identified difficulties with the limited availability of these specialists OOH whilst trying to run a 24-hour service in the department and the need to make a quick decision. During OOH, it was identified that admission avoidance became a difficulty due to a lack of resources for the older person both within the ED and in the community as these specialist staff are not on duty. OOH discharge was not deemed to be as safe as office hours. Often older adults would end up having to stay in ED or be admitted due to an identified risky discharge OOH, especially at weekends.

> “There is a huge differential in you coming and being seen by a Frailty Intervention Team during office hours on Monday to Friday and you coming to the emergency department at any other time.” (Consultant in Emergency Medicine No.1)

Respondents also reported a level of risk based on the variances of when assessments were undertaken, and their suitability for assessing older people attending the ED. This included triage and frailty assessments following ED presentation. Variances existed in relation to if, and when, these assessments took place, and the scales used. Some participants reported that frailty assessments were not undertaken at the initial triage assessment and not at all during OOH due to demands on and availability of specialist older person team members, and the numbers of attendees to the ED. Others reported it was the first important step and always tried to get older people assessed as quickly as possible. Again, variances occurred between office hours and OOH practice.

> “We do the clinical frailty scale…. The triage nurses are overwhelmed…. They can’t keep up with the tsunami of patients coming through, so that would be done post triage.” (Consultant in Emergency Medicine No. 1)

Several respondents reported strategies to reduce risk and ensure safety for older people when being discharged home OOH from the ED. These included arranging transport, providing a staff escort for support and to ensure safety, and using a discharge lounge which was only available during office hours. Practices varied between hospitals. Many hospitals witnessed changing variances in staff profiles and skill mix with staff turnover being significant in some areas. This change in personnel following the employment of many international nurses to Ireland was reported to be a difficulty for the older adult in the ED, with cultural and language differences evident. This was identified as a specific issue OOH when no specialist frailty team support was available to support these new international staff members who may be unfamiliar with the specific needs of older people on discharge from the ED. Thus, adding to a potential risk on OOH discharge. Other challenges identified included the increasing numbers of international older adults with growing health care needs living in inappropriate places such as hotels, and associated difficulties in accessing support on discharge from the ED.

> “There are issues with new staff from outside of Ireland, and the biggest problem is communication particularly for the elderly patient. Language is a big thing, same with the doctor. They just have a different culture. They just presume that there is going to be somebody at home to look after that patient, because that’s what they would do.” (Clinical Education Facilitator No. 4)

### Bright lights and noise

Participants identified several risks for the older adults who were delayed or detained in the ED OOH whilst awaiting discharge, often because of an overstimulated environment resulting in adverse consequences. These included significant biological and psychological effects such as sleep deprivation, anxiety, delirium, and medication and mobility issues. Older people also faced challenges within the ED environment, including being exposed to frightening incidents, and the risks of acquiring infections and pressure ulcers. A drop in the level of supervision and care from HCPs was also a concern once an older person was deemed fit for discharge but awaiting review by the specialist team.

> “Older people staying in an ED can…. get sicker, pressure sores and anxiety.…. you have to weigh up the pros and cons and what is safest in the long run and having to keep them in to be seen by the team the next morning.” (Clinical Nurse Specialist in Frailty, No. 2)

Keeping an older person in ED OOH for follow-up by the older person services although medically discharged was reported to result in an increase in patient experience times and a reduction in patient flow within the ED. This was identified as a cause of stress for staff and an increase in workload, due to overcrowding within the ED but also due to a reflection of poor ED performance because of prolonged patient experience times for those over 75 years of age.

> “Out of hours, where our colleagues notice an issue with a patient that would benefit from the multi-disciplinary team input, is when we would see that knock-on effect on time.” (Consultant in Emergency Medicine No.2)

It was reported that older people also faced challenges within the ED environment OOH, as often no specific older person area was available within the ED due to the volume of attendances, overcrowding and a lack of space. Respondents reported that this could lead to increased adverse outcomes for the older person because of difficulties in meeting fundamental needs, from having to stay overnight on a chair or trolley and often on a busy corridor.

> “…. for a patient who has poly morbidities, their risks of having a negative outcome are much higher. There is also the risk of them not getting appropriate medications, and meals as well as not getting the toilet ….” (Consultant in Emergency Medicine No.2)

### New ways of working

Participants interviewed recognised the difficulties in providing a 24-hour service in the ED, when their counterparts in other areas of the hospital and community services were not available. They were very forthcoming in providing recommendations for future practice to improve services for older people whilst being discharged from the ED OOH (see Figure 1). This included various aspects of discharge and care within the department and extending services to have specialist older person HCPs working 24/7 to ensure safe older person discharge from the ED OOH.

**Fig 1.**
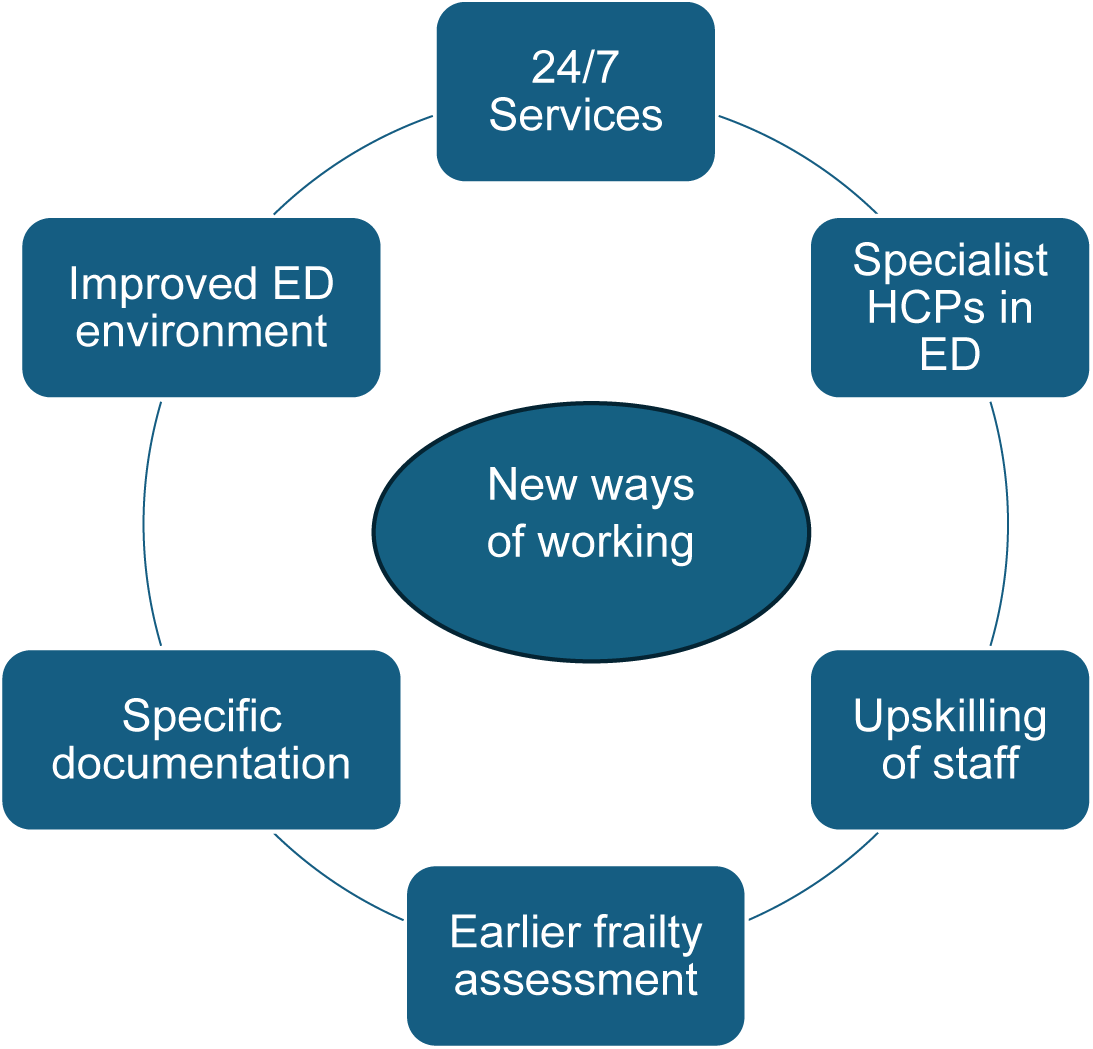
New ways of working-recommendations for practice.

Many respondents identified the need to improve experiences OOH. This included ED staffing OOH, and the need for more older person specialist health care professionals 24/7 in the ED including physicians in older person emergency medicine and employment of a nurse to ensure any required follow-up is organised for older people following ED discharge. Respondents also highlighted the need for upskilling of all ED staff. With the increasing older person demographics, it was identified that a need existed to ensure that ED staff had the knowledge and skills to be able to provide a 24/7 service safely and effectively for this specialist cohort. This could be in the form of a national education programme.

> “….. that the ED staff themselves need a greater skills set which could be provided by a 2-day course with online learning …. we wouldn’t contemplate having staff in the emergency department if they didn’t have ACLS or ATLS training and yet, there isn’t much of our work which is now cardiac arrest. There’s far more of our work that is older adult now.” (Consultant in Emergency Medicine No.1)

Participants in this study recommended a greater and earlier focus on frailty on a 24/7 basis, not just at triage in the ED, but also pre-attendance by ambulance personnel and at primary care centres. This was identified as being very important considering the growth in older people demographics internationally and the associated risk of an increased incidence of frailty.

> “Ideally frailty should be scored in triage from the minute the person presents to the door. I think everybody should be thinking frailty.…… to start scoring them coming through the door from triage. And even before that, the ambulance staff, the GP surgery.” (Clinical Nurse Specialist in Frailty No. 1)

Recommendations from participants also included a need to focus on specific documentation which could identify and highlight the risk of frailty for the older person presenting to the ED. It was suggested that a national discharge checklist could help to ensure that there is uniformity in discharging the older person from the ED. A focus on written instructions and patient education for older people in the ED was also suggested in order to have a record of treatment provided and further care required. Improvements to the ED environment were also suggested. These included specific frailty units and separate treatment areas for older people as seen internationally. It was suggested that these could help reduce the risk of adverse reactions for older adults presenting to the ED by the avoidance of frightening experiences in a busy, highly stimulated ED.

> “What I would say needs to happen is to have an older person’s unit, like a frailty unit part of the ED. In America some of the hospitals are just older people EDs because that’s the population they’re getting. Their whole focus in the 24 hours is older people. And that’s the way it’s going to be. It’s not going to go away.” (Clinical Nurse Specialist in Frailty, No. 1)

## Discussion

This paper identified the safety and risk aspects of HCP’s experiences of discharging older people during OOH versus office hours from the Emergency Department. Patient safety is defined as “the reduction of risk of unnecessary harm associated with healthcare to an acceptable minimum” (22). However, it is estimated that around one in ten patients are harmed during health care, resulting in substantial consequences for patients (23). Discharge processes are an important aspect of a patient’s health care journey. Many older adults who attend the ED for assessment are subsequently discharged. Participants in this study highlighted risks for both discharging older people home OOH without organised supports and in keeping older patients in the ED until services are available. The results identified important but diverse practices regarding older people ED discharge occurring in different geographical areas and between OOH and office hours. Despite its unique and challenging clinical environment, the principles of patient-centred care and shared decision making are evident within the ED (24) with a holistic problem-solving, person-centred approach to discharge needed (14). Participants in this study were cognisant of the wishes of the older person; however, this was alongside their concern over patient safety if an adverse event were to occur post discharge or indeed in the ED OOH whilst waiting until further assessment could take place during office hours.

Whilst some developments in ED care are not new, such as an admission avoidance (AA) team (25), in more recent years many welcome developments have been realised in older person ED care and follow up in the community. These aim to expand community healthcare services to be closer to where older people reside and to reduce the demand on hospital services by admission avoidance and transition from hospital to home (26), recognising the risks of hospitalisation for this cohort. A study exploring the impact of Home FIRsT (Frailty Intervention & Response Team) interventions on older patients’ experience times in the ED (27), identified a need for further research in the area. The authors recognised that assessments by the Home FIRsT can be lengthy and in EDs where there are waiting criteria (e.g. 4-hour National Health Service target), it would be particularly challenging to run such a service. Although the study excluded patients attending out of hours, the authors did suggest a future potential to deliver continuous Home FIRsT-like services 24 hours a day, seven days a week, both in the ED and in community services. Interestingly, ED PLUS, an initiative introduced to link the transition of care between the ED visit and the community, was shown to result in a lower incidence of functional decline and improvement in quality of life for the older adult post ED (28). Whilst these developments are very welcome, the findings in this study showed that many older person specialist teams are inaccessible in the ED or in the community when discharge occurs OOH which impacted on the decision to discharge.

Point-of-care identification is needed for older adults presenting to the ED who may be at risk of adverse outcomes (29). Assessments including triage, comprehensive geriatric assessment, frailty and delirium assessments are regularly undertaken with the older person attending the ED. Zachariasse et al (30) and Brouns et al (31) reported that the validity of the Manchester Triage System in emergency care was moderate to good; however, lowest performance was in older patients over 65 years of age. Questions over the use of the Manchester Triage System with older adults in the ED also emerged in this study, with some participants expressing concerns over its suitability. Potential risks for older adults with frailty in the ED have been recognised. Frailty is often assessed using the Clinical Frailty Scale on a scale from 1-9, assessing domains such as comorbidity, function, and cognition (32). Significant findings from a cross-sectional study in 14 European countries, identified that 40% of older people attending the ED had a score of 5 or more when assessed using Rockwood et al’s (33) Version 1.2 Clinical Frailty Scale (34). The use of a clinical frailty scale in ED may assist with prompt decision making at triage (35), timely access to appropriate care for older patients attending (36; 37), and in obtaining a more holistic clinical assessment (38). Additionally, early assessment and intervention in ED can increase patient satisfaction levels (5). Although not OOH focused, O’Shaughnessy et al (39), also identified that comprehensive geriatric assessment may help improve patient outcomes by achieving admission avoidance and thus avoiding any possible adverse consequences. However, the ED environment is not appropriate for performing such in-depth assessments, with a need for quick and user-friendly tools (40) as frailty screening should be undertaken within four hours of attending the ED and take less than five minutes to complete (41). Daily encounters with older people with frailty will be a norm for almost all health and social care professionals (42). Therefore, staff must be cognisant of increased frailty especially during long department waits in overcrowded and sensory heightened areas present in many EDs. Despite the reported benefits of using a Clinical Frailty Scale at triage, this study identified that frailty assessment is not performed at triage in some hospitals and often never undertaken OOH, resulting in delayed or missed identification of some older adults with frailty.

Whilst early ED frailty screening at triage can transform care through fast tracking to required care pathways, education for all professionals is essential to ensure engagement in and recognition of frailty (43). Despite the prevalence of frailty, there is a lack of training in medical and nursing curricula and a need exists for education and skills development for HCPs (44). Emergency services must be equipped to provide care for older patients who present with complex health and care needs (34). Assessments can also be compounded by language barriers in nurse/patient communication which Ali and Watson (45) reported can have adverse effects on the provision of appropriate and effective care. Indeed, the World Health Organisation (46) highlighted the increase in the international migration and mobility of health workers. In this present study, participants identified communication issues between staff and older patients, and cultural and decision-making concerns during older person ED discharge OOH. Health education is an important aspect of ensuring safe discharge re treatment and follow-up. However, participants reported it as a difficulty for many patients being discharged from the ED. A need exists to ensure a greater adherence to discharge instructions and to improve the transition from ED to home (47), possibly using a multimodal approach in delivering discharge instructions (48). Participants in this study highlighted issues surrounding ED discharge information for older adults and recommended the need for specific older patient discharge information leaflets.

ED discharge is associated with increased risk with up to six months decline in functional status for the older adult living in the community (49). Concerningly almost 80% of older people are discharged from the ED with a minimum of one unaddressed health concern (Memedovich et al (50). Condon et al (51) explored the transition of older adults to the community from the ED. Whilst the qualitative review was not OOH focused, findings identified fragmented care, communication difficulties, reports of unplanned discharges with transport not being organised for older adults. Participants in this study identified that they had to consider many factors to ensure a safe older person ED discharge including time, distance, and transport issues. Discharging an older adult living alone in a rural area may not be a satisfactory start to the discharge process. In Ireland, around 5% of the population live 50km or more from an adult emergency department while 0.8% of the population have a travel distance of 75km or more (52). Time of discharge can be challenging for older people, especially at night when support networks and services in the community are less available (14).

This study also highlighted the risks in being detained in ED whilst awaiting further assessments prior to discharge. Older person ED discharge needs to be safe and effective on a 24/7 basis. However, although not specific to OOH, some of the challenges caring for older people in the ED, included the unsuitable physical environment and equipment, communication issues, a limited time factor, and a lack of geriatric-specific knowledge (53). Attendance at an ED can have adverse consequences for the older patient with dementia (54; 8), which can be very distressing but avoidable for patients and their families. Healthcare systems benefit from providers delivering a positive patient experience (55). However, long waits, difficult waiting environments, lack of communication, privacy, and personal care have been identified previously as areas of concern for older people in the ED (7). Additionally, many healthcare needs of older adults are not being met either in the ED or by interventions initiated by the ED (50). Similarly, in this study, participants recognised the potentially serious risks in keeping an older adult for extended periods of time in the ED, which included risk of infection, delirium, anxiety, and possible neglect. Often this delay OOH was due to the perceived risks for the older person, and the unavailability of specialist staff who only work during office hours. This is a major area of concern as these serious risks are preventable if adequate resources were in place.

ED overcrowding results in decreased access to an appropriate treatment space, and a decrease in adherence to the National Emergency Access Target (NEAT) (56). An increase in older people attendances with complex and chronic conditions are causes of overcrowding and it is recognised that there is need for efficient ED patient flow, using initiatives to meet timed patient disposition targets, as well as expanded primary care (57). Internationally, older adults have advocated for older adult services such as a geriatric ED to meet specific health needs for their cohort which could help to reduce overcrowding and wait times (58; 7. Similar difficulties were also highlighted in this study including increased wait times and older people having to frequently stay overnight on a chair in an over stimulated ED corridor, due to overcrowding, with a recommendation for specific older person areas.

Lowthian et al (59) reported a need for further research around safe older person ED discharge as a lack of consensus existed as to what forms evidence-based best practice for patients discharged from the ED. Enhancement of integrated care is required especially with regards to improved communications, sharing of responsibility between care providers, additional resources and further research evaluating care practices (60). Recognising the need to improve care for older patients in the ED, Lucke et al (61) developed expert clinical recommendations for Geriatric Emergency Medicine (GEM). Mooijaart et al (62) also recognised the rapid development of GEM in response to ageing demographics, proposing that further work, research and innovation is needed to provide holistic care for the older adult, including organisation of care, identification of patient preferences and needs, and improving HCP’s knowledge and expertise. Respondents in this study also identified a need for improved documentation, staff training, and national discharge guidelines to ensure uniformity of discharge practices for older people from all EDs on a 24/7 basis.

### Strengths and weaknesses

This study had several strengths and weaknesses. Firstly, whilst considerable research has been undertaken on ED matters in recent years, a dearth still exists in relation to out of hours ED discharge experiences. This study is the first of its kind to explore this important topic and has addressed the gap in research which existed. It is likely that the practices, experiences, and concerns regarding out of hours discharge seen in this study also pertain to EDs globally although there is a need for further international research. Secondly, participants included HCPs from a variety of backgrounds and from a wide geographical spread which enhanced the rigour of the study. This study identified diverse practices in healthcare, taking cognisance of the availability of resources and follow-up care, the geographical landscape, demographics, and cultural aspects in caring for older family members. However, additional participants such as occupational therapists and social workers may have generated additional insightful experiences.

### Conclusions

Whilst the older adult cohort are one of the main attenders at EDs, many are subsequently discharged home. However, this study highlighted that if deemed not for admission but unfit for discharge home, older adults may experience a delayed discharge OOH. This principally occurs if a perceived biological/psychological/social risk is identified and/or if further assessment is required by members of the multi-disciplinary team, yet the service is unavailable OOH. Due to the multiple contesting factors within a dynamic emergency department, attendance and long waits therein can lead to a risk of serious adverse effects for the older adult including discomfort from inappropriate waiting areas, a lack of sleep, anxiety, and delirium. Patient experience times and ED flow can also be impacted. To ensure a safe and effective discharge home 24/7 in every ED regardless of location, there is a need to address gaps in care identified. With the significant increase in older person demographics, there is an urgent need for healthcare managers and relevant policy makers to develop best practice guidelines, ensure greater availability of older people services and teams both in the ED and in the community, increase explicit older person ED treatment areas, and develop specific education courses and upskilling for ED staff. These recommendations can assist in achieving a safe and appropriate older person ED discharge around the clock.

## Data Availability

All data underlying the results presented in the study cannot be shared publicly due to ethical restrictions and the personal nature of the responses which could be identifiable in their raw state. All data will be available elsewhere with limitations from the Ulster University Research Governance Department. Ms. Elaine Bell, Research Governance Officer Phone: +44 28 9536 5028

## Supporting information

S1 Appendix 1: Interview schedule (DOC)

S2 Appendix 2 Standards for reporting qualitative research checklist (SRQR) (DOC)

## Acknowledgements

The authors acknowledge the participating health care professionals for their interest, time, and honesty in this study, along with support and guidance received from members of the National Clinical Programme for Emergency Medicine and the Irish Association for Emergency Medicine.

## Conflict of interest

The authors declare no conflict of interest.

## Author Contributions

**Conceptualization:** Mary Dunnion, Assumpta Ryan, Debbie Goode, Sonja McIlfatrick.

**Formal analysis:** Mary Dunnion, Assumpta Ryan, Debbie Goode, Sonja McIlfatrick.

**Investigation:** Mary Dunnion.

**Methodology:** Mary Dunnion, Assumpta Ryan, Debbie Goode, Sonja McIlfatrick.

**Project administration:** Mary Dunnion.

**Supervision:** Assumpta Ryan, Debbie Goode, Sonja McIlfatrick.

**Writing– original draft:** Mary Dunnion

**Writing– review & editing:** Mary Dunnion, Assumpta Ryan, Debbie Goode, Sonja McIlfatrick.

## Notes

### Competing Interest Statement

The authors have declared no competing interest.

### Funding Statement

The author(s) received no specific funding for this work.

### Author Declarations

Ethical approval was obtained from Ulster University Research Ethics Filter committee (Project Number: FCNUR-22-017) as well as from the Research Ethics Committees (RECs) in the hospitals involved in the study.

## References

1. World Health Organisation. Ageing and health. 2022. Available: https://www.who.int/news-room/fact-sheets/detail/ageing-andhealth#:~:text=Every%20country%20in%20the%20world%20is%20experiencing%20growth,from%201%20billion%20in%202020%20to%201.4%20billion. Accessed: 07/06/2024

2. United Nations General Assembly. United Nations Decade of Healthy Ageing (2021-2030): resolution/adopted by the General Assembly. 2020. United Nations, New York. Available: https://policycommons.net/artifacts/8967746/united-nations-decade-of-healthy-ageing-2021-2030/9835673/ Accessed: 07/06/2024.

3. Eliopoulos C. Gerontological Nursing (Tenth edition). China, Wolters Kluwer; 2021.

4. World Health Organization. Decade of Healthy Ageing Baseline Report. World Health Organization, Geneva; 2020.

5. Cassarino M, Robinson K, Trépel D, O’Shaughnessy Í, Smalle E, White S, Devlin C, Quinn R, Boland F, Ward ME. and McNamara R. Impact of assessment and intervention by a health and social care professional team in the emergency department on the quality, safety, and clinical effectiveness of care for older adults: A randomised controlled trial. PLoS Medicine, 2021; 18(7), p.e1003711.

6. Corcoran G, Bernard P, Kenna L, Malone A, Horgan F, O’Brien C, Ward P, Howard W, Hogan L, Mooney, R., Masterson, S. “Older People Want to Be in Their Own Homes”: A Service Evaluation of Patient and Carer Feedback after Pathfinder Responded to Their Emergency Calls. Prehospital Emergency Care. 2023;27(7):866–874.

7. Mwakilasa, M. T., Foley, C., O’Carroll, T., Flynn, R., & Rohde, D. Care experiences of older people in the emergency department: a concurrent mixed-methods study. Journal of Patient Experience, 2021; 8, doi:10.1177/23743735211065267.

8. Goode, D., Ryan, A., Melby, V., & Slater, P. Care experiences of older people with mental health needs and their families in emergency medical services settings. International Journal of Older People Nursing, 2023; 18, e12500. 10.1111/opn.12500

9. Rowe, A., & Knox, M. The impact of the healthcare environment on patient experience in the Emergency Department: a systematic review to understand the implications for patient-centered design. HERD: Health Environments Research & Design Journal, 2023; 16(2), 310–329.

10. Greenwald, P.W., Estevez, R.M., Clark, S., Stern, M.E., Rosen, T., Flomenbaum, N. The ED as the primary source of hospital admission for older (but not younger) adults. The American Journal of Emergency Medicine, 2016, 34, 6, pgs. 943–947. ISSN 0735-6757. 10.1016/j.ajem.2015.05.041

11. Cetin-Sahin D, Ducharme F, McCusker J, Veillette, N., Cossette, S., Minh Vu, T., Vadeboncoeur, A., Lachance, P., Mah, R., Berthelot, S. Experiences of an Emergency Department Visit Among Older Adults and Their Families: Qualitative Findings from a Mixed-Methods Study. Journal of Patient Experience, 2020; 7(3):346–356. doi:10.1177/2374373519837238

12. Gettel, C.J., Serina, P.T., Uzamere, I., Hernandez-Bigos, K., Venkatesh, A.K., Rising, K.L., Goldberg, E.M., Feder, S.L. Cohen, A.B., Hwang, U. Emergency department-to-community care transition barriers: A qualitative study of older adults. J Am Geriatr Soc., 2022; 70(11): 3152–3162.

13. Dunnion, M., Ryan, A., Goode, D., McIlfatrick, S. Supporting older people following out of hours discharge from the Emergency Department: An integrative review of the literature. International Journal of Older People Nursing, 2023; 18(3): e12529.

14. Hyslop, B. ‘Not safe for discharge’? Words, values, and person-centred care. Age and Ageing, 2020; 49(3), 334–336.

15. World Health Organisation. World report on ageing and health. Geneva: World Health Organization.2015.Available:https://iris.who.int/bitstream/handle/10665/326843/WHO-FWC-ALC-19.1-eng.pdf?sequence=17 Accessed: 19/08/2024.

16. Integrated Care Programme for Older People Steering Group. Making a start in integrated care for older persons a practical guide to the local implementation of integrated care programmes for older persons. 2017. Available: https://www.hse.ie/eng/services/publications/clinical-strategy-and-programmes/a-practical-guide-to-the-local-implementation-of-integrated-care-programmes-for-older-persons.pdf Accessed 22/7/2024.

17. Braun, V. & Clarke, V. Thematic Analysis. A Practical Guide. Sage Publications; 2022.

18. Braun, V. & Clarke, V. One size fits all? What counts as quality practice in (reflexive) thematic analysis? Qualitative Research in Psychology, 2021; 18:3, 328–352.

19. Jamieson, M. K., Govaart, G. H., & Pownall, M. Reflexivity in quantitative research: A rationale and beginner’s guide. Social and Personality Psychology Compass, 2023; 17(4), e12735.

20. Olmos-Vega, F. M., Stalmeijer, R. E., Varpio, L., & Kahlke, R. A practical guide to reflexivity in qualitative research: AMEE Guide No. 149. Medical teacher, 2023; 45(3), 241–251.

21. O’Brien BC, Harris IB, Beckman TJ, Reed DA, Cook DA. Standards for reporting qualitative research: a synthesis of recommendations. Academic Medicine, 2014; 89 (9):1245–1251. DOI: 10.1097/ACM.0000000000000388

22. World Health Organisation Patient Safety and World Health Organization. Conceptual framework for the international classification for patient safety version 1.1: final technical report January 2009 (No. WHO/IER/PSP/2010.2). Report No.: 606940937X. Geneva, World Health Organization, 2010.

23. Slawomirski, L., Auraaen, A., Klazinga, N.S. “The economics of patient safety: Strengthening a value-based approach to reducing patient harm at national level”, OECD Health Working Papers, 2017, No. 96, OECD Publishing, Paris. 10.1787/5a9858cd-en.

24. Hess, E. P., Grudzen, C. R., Thomson, R., Raja, A. S., & Carpenter, C. R. Shared decision-making in the emergency department: respecting patient autonomy when seconds count. Academic Emergency Medicine, 2015; 22(7), 856–864.

25. Hardy C, Whitwell D, Sarsfield B, Maimaris, C. Admission avoidance and early discharge of acute hospital admissions: an accident and emergency-based scheme. Emergency Medicine Journal, 2001; 18:435–440.

26. Health Service Executive. Enhanced Community Care. 2024. Available: https://www.hse.ie/eng/services/list/2/primarycare/enhanced-community-care/#:~:text=The%20ECC%20programme%20has%20delivered:%201.%2096%20of Accessed 17/07/2024.

27. O’Shaughnessy, Í., Romero-Ortuno, R., Edge, L., Dillon, A., Flynn, S., Briggs, R., Shields, D., McMahon, G., Hennessy, A., Kennedy, U. and Staunton, P. Home FIRsT: interdisciplinary geriatric assessment and disposition outcomes in the emergency department. European journal of internal medicine, 2021; 85, pp.50–55.

28. Conneely, M., Leahy, S., O’Connor, M., Corey, G., Gabr, A., Saleh, A., Okpaje, B., O’Shaughnessy, Í., Synnott, A., McCarthy, A. and Holmes, A. A physiotherapy-led transition to home intervention for older adults following emergency department discharge: a pilot feasibility randomised controlled trial (ED PLUS). Clinical Interventions in Aging, 2023; pp.1769–1788.

29. Carpenter, C. R., Shelton, E., Fowler, S., Suffoletto, B., Platts-Mills, T. F., Rothman, R. E., & Hogan, T. M. Risk factors and screening instruments to predict adverse outcomes for undifferentiated older emergency department patients: a systematic review and meta-analysis. Academic Emergency Medicine, 2015; 22(1), 1–21.

30. Zachariasse, J.M., Seiger, N., Rood, P.P., Alves, C.F., Freitas, P., Smit, F.J., Roukema, G.R. and Moll, H.A. Validity of the Manchester Triage System in emergency care: A prospective observational study. PloS ONE, 2017; 12(2), p.e0170811.

31. Brouns, S. H., Mignot-Evers, L., Derkx, F., Lambooij, S. L., Dieleman, J. P., & Haak, H. R. Performance of the Manchester triage system in older emergency department patients: a retrospective cohort study. BMC Emergency Medicine, 2019; 19, 1–11.

32. Church, S., Rogers, E., Rockwood, K., & Theou, O. A scoping review of the Clinical Frailty Scale. BMC geriatrics, 2020; 20, 1–18.

33. Rockwood, K., Song, X., MacKnight, C., Bergman, H., Hogan, D.B., McDowell, I., Mitnitski, A. A global clinical measure of fitness and frailty in elderly people. Canadian Medical Association Journal, 2005; 173(5):489–495.

34. European Taskforce on Geriatric Emergency Medicine (ETGEM) Collaborators. Prevalence of Frailty in European Emergency Departments (FEED): an international flash mob study. European Geriatric Medicine, 2024; 15(2):463–470.

35. Elliott, A., Taub, N., Banerjee, J., Aijaz, F., Jones, W., Teece, L., van Oppen, J. and Conroy, S. Does the clinical frailty scale at triage predict outcomes from emergency care for older people? Annals of emergency medicine, 2021; 77(6), pp.620–627.

36. Blomaard, L.C., Speksnijder, C., Lucke, J.A., de Gelder, J., Anten, S., Schuit, S.C., Steyerberg, E.W., Gussekloo, J., de Groot, B. and Mooijaart, S.P. Geriatric screening, triage urgency, and 30-day mortality in older emergency department patients. Journal of the American Geriatrics Society, 2020; 68(8), pp.1755–1762.

37. Ng, C.J., Chien, L.T., Huang, C.H., Chaou, C.H., Gao, S.Y., Sherry, Y.H.C., Hsu, K.H. & Chien, C.Y. Integrating the clinical frailty scale with emergency department triage systems for elder patients: A prospective study. The American Journal of Emergency Medicine, 2023; 66, pp.16–21.

38. Elliott, A., Phelps, K., Regen, E., & Conroy, S. P. Identifying frailty in the Emergency Department—feasibility study. Age and ageing, 2017; 46(5), 840–845.

39. O’Shaughnessy, Í., Robinson, K., Whiston, A., Barry, L., Corey, G., Devlin, C., Hartigan, D., Synnott, A., McCarthy, A., Moriarty, E. and Jones, B. Comprehensive geriatric assessment in the emergency department: a prospective cohort study of process, clinical, and patient-reported outcomes. Clinical Interventions in Aging, 2024; pp.189–201.

40. van Dam, C.S., Hoogendijk, E.O., Mooijaart, S.P., Smulders, Y.M., de Vet, R.C., Lucke, J.A., Blomaard, L.C., Otten, R.H., Muller, M., Nanayakkara, P.W. and Trappenburg, M.C. A narrative review of frailty assessment in older patients at the emergency department. European Journal of Emergency Medicine, 2021; 28(4), pp.266–276.

41. Moloney, E., O’Donovan, M.R., Carpenter, C.R., Salvi, F., Dent, E., Mooijaart, S., Hoogendijk, E.O., Woo, J., Morley, J., Hubbard, R.E. and Cesari, M. Core requirements of frailty screening in the emergency department: an international Delphi consensus study. Age and ageing, 2024; 53(2), p. afae013.

42. Conroy, S., Carpenter, C., Banerjee, J. Silver Book II Quality Care for Older People with Urgent Care Needs [Online]. 2021. Available: Silver Book II | British Geriatrics Society (bgs.org.uk) Accessed: 07/06/2024.

43. van Oppen, J. D., & Suzanne, M. Frailty screening in the Emergency Department: why does it matter? Age and Ageing, 2024; 53(4), afae056.

44. Hammel, I. S., & Chapman, E. N. Frailty training among healthcare professionals. The Journal of nutrition, health and aging, 2024; 28 (6), 100258.

45. Ali, P. A., & Watson, R. Language barriers and their impact on provision of care to patients with limited English proficiency: Nurses’ perspectives. Journal of Clinical Nursing, 2018; 27(5-6), e1152–e1160.

46. World Health Organisation. Addressing the international migration of health workers. 2024. Available: Addressing the international migration of health workers (who.int) Accessed: 13/8/2024.

47. Benjenk, I., DuGoff, E.H., Jacobsohn, G.C., Cayenne, N., Jones, C.M., Caprio, T.V., Cushman, J.T., Green, R.K., Kind, A.J., Lohmeier, M. and Mi, R. Predictors of older adult adherence with emergency department discharge instructions. Academic Emergency Medicine, 2021; 28(2), pp.215–225.

48. Hoek, A. E., Anker, S. C., van Beeck, E. F., Burdorf, A., Rood, P. P., & Haagsma, J. A. Patient discharge instructions in the emergency department and their effects on comprehension and recall of discharge instructions: a systematic review and meta-analysis. Annals of Emergency Medicine, 2020; 75(3), 435–444.

49. Nagurney, J. M., Fleischman, W., Han, L., Leo-Summers, L., Allore, H. G., & Gill, T. M. Emergency department visits without hospitalization are associated with functional decline in older persons. Annals of emergency medicine, 2017; 69(4), 426–433.

50. Memedovich, A., Asante, B., Khan, M., Eze, N., Holroyd, B. R., Lang, E., Kashuba, S., Clement, F. Strategies for improving ED-related outcomes of older adults who seek care in emergency departments: a systematic review. International Journal of Emergency Medicine, 2024; 17, 16. 10.1186/s12245-024-00584-7.

51. Condon, B., Griffin, A., Fitzgerald, C., Shanahan, E., Glynn, L., O’Connor, M., Hayes, C., Manning, M., Galvin, R., Leahy, A. and Robinson, K. Older adults experience of transition to the community from the emergency department: a qualitative evidence synthesis. BMC geriatrics, 2024; 24(1), p.233.

52. Central Statistics Office. Measuring Distance to Everyday Services in Ireland [Health Services Measuring Distance to Everyday Services in Ireland - Central Statistics Office](https://www.cso.ie/en/releasesandpublications/ep/p-mdsi/measuringdistancetoeverydayservicesinireland/healthservices/). 2024. Available: [Introduction Measuring Distance to Everyday Services in Ireland - Central Statistics Office](https://www.cso.ie/en/releasesandpublications/ep/p-mdsi/measuringdistancetoeverydayservicesinireland/introduction/) Accessed 19/07/202

53. Lennox, A., Braaf, S., Smit, D. V., Cameron, P., & Lowthian, J. A. (2019) Caring for older patients in the emergency department: health professionals’ perspectives from Australia–the safe elderly emergency discharge project. Emergency Medicine Australasia, 2019; 31(1), 83–89.

54. Watkins, S., Murphy, F., Kennedy, C., Graham, M., & Dewar, B. Caring for older people with dementia in the emergency department. British Journal of Nursing, 2020; 29(12), 692–699.

55. Aleksandrovskiy, I., Ganti, L., Simmons, S. The Emergency Department Patient Experience: In Their Own Words. Journal of Patient Experience, 2022; 20;9,1–4.

56. Smith, B., Bouchoucha, S., & Watt, E. ‘Care in a chair’–The impact of an overcrowded Emergency Department on the time to treatment and length of stay of self-presenting patients with abdominal pain. International Emergency Nursing, 2016; 29, 9–14.

57. Morley, C., Unwin, M., Peterson, G. M., Stankovich, J., & Kinsman, L. Emergency department crowding: a systematic review of causes, consequences, and solutions. PloS ONE, 2018; 13(8), e0203316.

58. American College of Emergency Physicians. Policy Statement. Safe Discharge from the Emergency Department. Dallas, American College of Emergency Physicians. 2019.

59. Lowthian, J. A., McGinnes, R. A., Brand, C. A., Barker, A. L., & Cameron, P. A. Discharging older patients from the emergency department effectively: a systematic review and meta-analysis. Age and ageing, 2015; 44(5), 761–770.

60. Dunlea, S., McCombe, G., Broughan, J., Carroll, Á., Fawsitt, R., Gallagher, J., Melin, K. and Cullen, W. Priorities in integrating primary and secondary care: a multimethod study of GPs. Journal of Integrated Care, 2022; 31(5), pp.1–14.

61. Lucke JA, Mooijaart SP, Heeren P, Singler K, McNamara R, Gilbert T, Nickel CH, Castejon S, Mitchell A, Mezera V, Van der Linden L, Lim SE, Thaur A, Karamercan MA, Blomaard LC, Dundar ZD, Chueng KY, Islam F, de Groot B, Conroy S. Providing care for older adults in the Emergency Department: expert clinical recommendations from the European Task Force on Geriatric Emergency Medicine. European Geriatric Medicine, 2022; 13(2):309–317. doi: 10.1007/s41999-021-00578-1. Epub 2021 Nov 5. PMID: 34738224; PMCID: PMC8568564.

62. Mooijaart, S. P., Carpenter, C. R., & Conroy, S. P. Geriatric emergency medicine—a model for frailty friendly healthcare. Age and Ageing, 2022; 51(3), afab280.

